# *In Vitro* Nasal Tissue Model for the Validation of Nasopharyngeal and Mid-turbinate Swabs for SARS-CoV-2 Testing

**DOI:** 10.1101/2021.11.22.21266713

**Authors:** Devon R. Hartigan, Miryam Adelfio, Molly E. Shutt, Stephanie M. Jones, Shreya Patel, Joshua T. Marchand, Pamela D. McGuinness, Bryan O. Buchholz, Chiara E. Ghezzi

## Abstract

Large-scale population testing is a key tool to mitigate the spread of respiratory pathogens, as in the current COVID-19 pandemic, where swabs are used to collect samples in the upper airways (e.g. nasopharyngeal and mid-turbinate nasal cavities) for diagnostics. However, the high volume of supplies required to achieve large-scale population testing has posed unprecedented challenges for swab manufacturing and distribution, resulting in a global shortage that has heavily impacted testing capacity world-wide and prompted the development of new swabs suitable for large-scale production. Newly designed swabs require rigorous pre-clinical and clinical validation studies that are costly and time consuming (*i*.*e*. months to years long); reducing the risks associated with swab validation is therefore paramount for their rapid deployment. To address these shortages, we developed a 3D-printed tissue model that mimics the nasopharyngeal and mid-turbinate nasal cavities, and we validated its use as a new tool to rapidly test swab performance. In addition to the nasal architecture, the tissue model mimics the soft nasal tissue with a silk-based sponge lining, and the physiological nasal fluid with asymptomatic and symptomatic viscosities of synthetic mucus. We performed several assays comparing standard flocked and injection-molded swabs. We quantified the swab pick-up and release, and determined the effect of viral load and mucus viscosity on swab efficacy by spiking the synthetic mucus with heat-inactivated SARS-CoV-2 virus. By molecular assays, we found that injected molded swabs performed similarly or superiorly in comparison to standard flocked swabs and we underscored a viscosity-dependent difference in cycle threshold values between the asymptomatic and symptomatic mucus for both swabs. To conclude, we developed an *in vitro* nasal tissue model, that corroborated previous swab performance data from clinical studies, with the potential of providing researchers with a clinically relevant, reproducible, safe, and cost-effective validation tool for the rapid development of newly designed swabs.

## Introduction

The rapidly increasing demand for COVID-19 testing since the start of the 2020 pandemic has caused significant bottlenecks in testing capacity due to a global shortage of testing supplies, including specimen collection swabs ^1-2^. To help overcome the swab shortage, alternative swabs, that could be mass produced at relatively low cost, *e*.*g*. via injection-molding processing, have been recently developed and raced to the market ^3^. Compared to a standard flocked, the injection-molded swabs are characterized by a non-absorbent head, and have demonstrated a more efficient release of viral RNA while absorbing less solution ^1, 4-5^. As a result, several prototypes of injection-molded swabs have been recently commercialized, including nasopharyngeal and mid-turbinate swabs like the IM2 and Rhinostic swabs ^3-4^. These new one-piece specimen collection swabs can be efficiently mass produced, without multistep manufacturing methods, and post-processing. Validation for swab prototypes typically requires preclinical testing before transitioning to clinical studies, which can take months to years. To streamline this initial pre-clinical validation process, there is compelling need for the development of an *in vitro* experimental model that recapitulates physical and structural features of the human nasal cavities to bridge bench-top and clinical studies ^5-10^. An *in vitro* nasal model can be efficiently and safely used by stakeholders to perform preclinical evaluations, anticipating device design modifications, before confidently moving to design-lock stage during clinical studies.

There are currently no tissue models available for this purpose. Bench top studies are performed by dipping in saline solutions without mimicking any of the physiological aspects of the nasal passage (i.e., architecture, mechanical and physical structures) nor of the actual swabbing procedure ^4, 11^. Other alternative modes of validation for prototype swabs in pre-clinical phase include swabbing the cheeks of participants and quantifying bacterial and cellular uptakes in comparison to the standard swabs ^12^ or through clinical studies ^3, 13^. By solely relying on clinical studies even for pre-clinical validation, the swabs meet a great deal of variability, thus needing to expand participant enrollment, with a significant increase in associated time and costs. Therefore, we hypothesize that the initial validation of swab prototypes on a simplified, reliable, and physiologically relevant nasal tissue model would provide more consistent and reproducible results, allowing investigators to assess swab performance in a time and cost-efficient manner ^5, 12^. Expanding the preclinical evaluation based on an *in vitro* tissue model will further support clinical studies to assess swab efficacy, streamlining the overall validation process.

Based on our previously developed anterior nasal tissue model ^5^, here we describe the design and fabrication of an *in vitro* tissue model platform (**Figure 1**) that aims to support pre-clinical validation of nasopharyngeal and mid-turbinate swabs, in an effort to significantly decrease swab validation time, allowing faster and more efficient distribution ^3^. The *in vitro* model is based on three dimensionally (3D)-printed nasal cavities to accurately mimic native tissue architecture, lined with a silk sponge to recapitulate the soft tissue structure. In addition, we varied viral load and mucus viscosities to better encompass the wide spectrum of clinical conditions, and further investigate their effects on swab performance.

**Figure 1.**
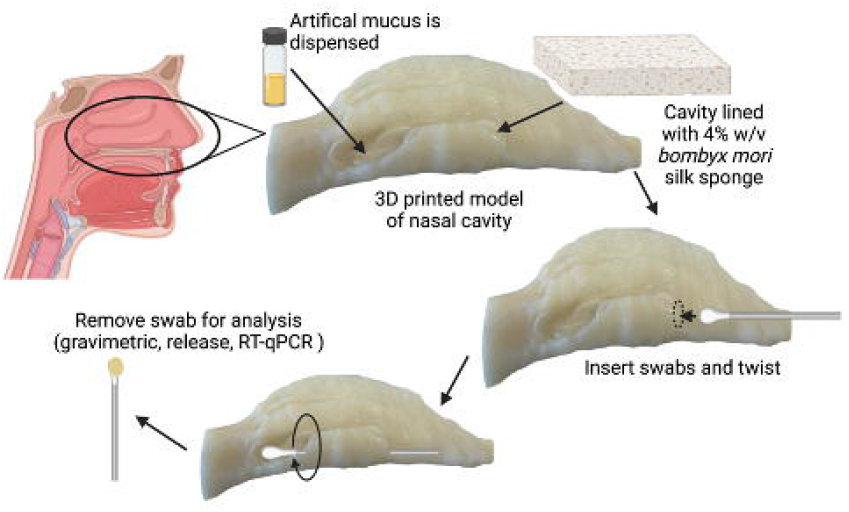
Nasal tissue benchtop model for swab validation. The 3D model replicates the architecture and structure of the human nasal cavity. 4% w/v silk sponges line the cavity, saturated with an artificial nasal mucus that physiologically mimics the viscosity of nasal fluid. A nasopharyngeal swab is inserted all the way until it meets resistance, while a mid-turbinate swab is inserted halfway. Both swabs are twisted and held for 15 seconds before they are removed. The swabs can be placed in diagnostic assay solutions and be ready for post collection analyses (PCR, gravimetric, release etc.). Created with BioRender.com.

To further support the use of the *in vitro* nasal model, we developed several validation assays to assess the performance of nasopharyngeal and mid-turbinate injection-molded and standard flocked swabs. We proposed new assessments to streamline initial swab validation, including gravimetric analysis and release quantification of fluorescently labelled microparticles, that mimic cellular material, to quantify swab pick-up and release capabilities. In addition, we carried out a Quantitative Reverse Transcription Polymerase Chain Reaction (RT-qPCR) using spiked mucus samples to mimic clinical swabbing and compare *in vitro* performance of the different types of swabs. The proposed model is a novel approach to support initial swab validation, as it accurately replicates the physiological components of the nasal cavity including architecture, structural elements, as well as physiologically relevant nasal fluids.

## Material and Methods

### Experimental Swabs

Herein, we assessed the performance of injection molded (IM) swabs in comparison to CLIA use approved Class I exempt standard flocked (SF) swabs. Obecare Sterile flocked Nasopharyngeal (NP) Swabs (Obecare, West Virginia) were used as the standard nasopharyngeal and mid-turbinate swabs (MT) in the experiments. The Obecare SF-NP swabs are standard flocked swabs characterized by an adhesive coated surface and nylon fibers that are attached perpendicularly for maximum absorbance (Obecare, West Virginia). IM-NP and IM-MT swabs were manufactured as a single element, based on a biocompatible polymer injected into a mold of a swab and allowed to harden (Yukon Medical, Durham, NC). (**Figure 2**).

**Figure 2.**
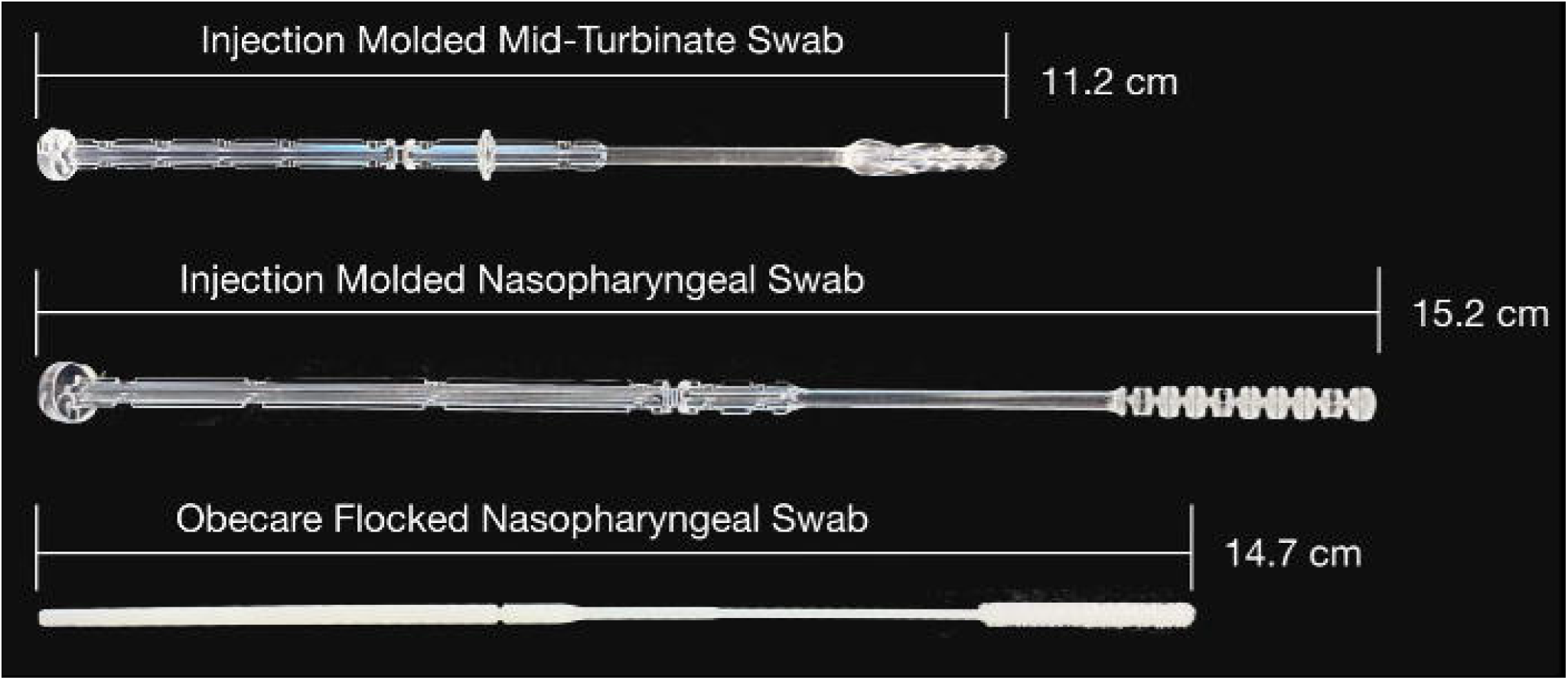
Macroimages of Mid-turbinate and Nasopharyngeal experimental swabs. Three swabs were used in this study, from top to bottom: 11.2 cm IM-MT swab, 15.7 cm IM-NP swab, and 14.7 cm SF-NP swab, was used as both a nasopharyngeal and mid-turbinate swab.

### 3D Printed Nasal Tissue Model Preparation

To provide a bench top validation system for experimental swabs, a 3D nasal tissue model was developed to mimic the human architecture and soft tissue properties (**Figure 1**). The physiological architecture was recreated by replicating the nasal cavity, specifically the opening to the cavity, the inferior nasal concha (inferior turbinate), the septum, the hard palate, the soft palate, and the nasopharynx. The 3D-model design was previously generated by the Aerosol Research Lab at Carleton University in Ottawa (Canada) ^14^. The 3D model of the nasal cavities was then generated using Acrylonitrile Butadiene Styrene (ABS, Gizmo Dorks LLC, Temple City, CA) filament with a Fused Deposition Modeling 3D printer (ABS-P430, Stratasys, Eden Prairie, Minnesota).

To mimic the soft tissue of the nasal cavities, aqueous silk sponges were prepared as previously reported ^15-16^. Briefly, pure silk fibroin was extracted from *Bombyx mori* cocoons by degumming the fibers in a sodium carbonate solution (0.02 M) (Sigma-Aldrich, St. Louis, Missouri) for 30 min to remove sericin. The degummed fibers were rinsed three times and dried overnight before the solubilization in 9.3M lithium bromide (Sigma-Aldrich, St. Louis, Missouri) for 2h at 60°C. The obtained solution was dialyzed for 3 days against DI water using a standard grade regenerated cellulose dialysis tubing (3.5 kD MWCO, Spectrum Labs Inc, Rancho Dominguez, California). The solution was then centrifuged to remove impurities. Subsequently, silk sponges were made according to the published protocol ^16^. 1.5 mL of silk solution (4% w/v) were poured into a 24 well plate (VWR Scientific, Radnor, Pennsylvania) and frozen for two cycles of 24h at −20°C and −80°C. The frozen plate was lyophilized for 3 days. The sponges were autoclaved at 121°C to induce the change in the secondary structure of the protein and induce water insolubility. Finally, sponges were cut into 0.5 mm thick slices with an *ad hoc* sample cutter. The 3D printed model cavities were then lined with silk sponges to mimic the native soft tissue.

### Synthetic asymptomatic and symptomatic mucus preparation and characterization

Two nasal mucus conditions were designed to mimic the viscosity of asymptomatic and symptomatic nasal fluid conditions ^17^. Poly (ethylene oxide) (PEO) (Sigma-Aldrich, St. Louis, Missouri. MW 1,000,000) was used to replicate these conditions, as previously reported ^18^. Upon preliminary investigations (data not reported here) and previous literature ^18^, PEO concentrations were chosen as 0.5% and 3.0% w/v for asymptomatic and symptomatic conditions, respectively.

Viscosity analysis was performed by using a dynamic viscometer (Brookfield Viscometer-Massachusetts) to identify the physiological values of the nasal mucus. Briefly, PEO solutions (0.5% and 3% w/v) were incubated for 30 minutes at 37°C. After the stabilization of the torque (equal or above 10%), 0.5 mL of PEO solutions (N=3 per condition) were loaded, while maintained at 37°C, and the analysis was carried out between 0.1 and 100 s^-1^.

### Swab Pick Up Quantification

To quantify swab uptake, the nasal tissue model was saturated with the physiologically relevant mucus solution and the following swabbing procedure was performed in accordance to CDC guidelines ^19^. NP swabs were inserted into the nasal cavity until resistance was encountered, while the MT swab was inserted to the midway point. Both swabs were twisted around the surfaces five times, held in place for 15 seconds, and then removed. Each swab was then placed into phosphate buffer solution (1x PBS) (VWR Scientific, Radnor, Pennsylvania) for further processing. In addition, we compared the swabbing workflow with the nasal tissue model (MODEL method) against the current gold-standard benchtop swab validation procedure ^1, 4, 11^, that comprises sequentially dipping swabs into tubes with relevant solutions (TUBE method).

The pick-up swab quantification was performed by gravimetric analysis for IM-MT and IM-NP swabs in comparison to the commercially available SF swab and the weight of each swab (N=5) was recorded before and after the MODEL or the TUBE methods. Results were reported as mass uptake, for three independent experiments.

### Swab Release Quantification

To quantify swab release, we carried out two independent investigations. In order to efficiently assess cellular material uptake, we loaded the synthetic mucuses with fluorescently labeled microparticles (10 µm) to mimic cellular particulates into the artificial nasal solution. FITC-labeled microparticles (Sigma-Aldrich, St. Louis, MO), based on melamine resin, were homogenously added to the 0.5 % and 3 % w/v PEO solution. The solution was then dispensed into the tissue model and allowed to saturate the silk sponges. The above-mentioned swabbing procedure was performed. Each swab (N=5) was then removed and placed in 1 ml volume of 1x PBS. 100 μL aliquots were taken in triplicate and analyzed with a SpectraMax M2 plate reader at 490 nm excitation and 525 nm emission. Fluorescence signal was then reported as an expression of cellular-mimicking uptake.

To further assess swab uptake and release of viral material, the nasal model was saturated with both nasal solutions spiked with heat-inactivated SARS-CoV-2, USA-WA1/2020 (NR-52286, BEI Resources, ATCC, USA), and the swabbing procedure was performed, as described above. To investigate the effect of viral load on swab performance, the mucus was spiked with three different concentrations of inactivated virus (10^7^, 10^6^, and 10^5^ copies/mL). After the procedure, each swab was removed and placed into a tube with 350 µl of 1x PBS. The vial with the swab was then vortexed for 30 sec. 5 µl from each sample was then tested to quantify the detection of SARS-CoV-2. To evaluate the presence of SARS-CoV-2, we performed the CDC 2019-Novel Coronavirus (2019-nCoV) Real-Time RT-PCR Diagnostic Panel (https://www.fda.gov/media/134922/download), per manufacturer instructions using the 2019-nCoV_N2 combined Primer/Probe mix with Quantabio Ultraplex One-Step RT-qPCR ToughMix. Amplification was performed following manufacturer instructions with a QuantStudio™ 5 Real-Time PCR System (Thermo Fisher Scientific, Waltham, MA, USA). The results for each swab (N=5) were reported as cycle threshold (Ct) value.

### Statistical Analysis

Statistical analysis was performed using a Student’s T-test (T-test) and Analysis of Variance (ANOVA) single factor with a p-value of < 0.05 using Origin(Pro), Version 2021b OriginLab Corporation, Northampton, MA, USA. T-test was performed when comparing paired IM to SF swabs in the gravimetric analysis, quantitative release, and RT-qPCR. ANOVA was performed to investigate the effect of swab type, mucus, and viral load.

## Results

### Physical characterization of asymptomatic and symptomatic nasal fluids

Viscosity analysis was performed to identify conditions for the symptomatic and asymptomatic physiological viscosities of the nasal mucus. The asymptomatic and symptomatic viscosities were mimicked by using PEO at 0.5% and 3% w/v, respectively. Both mucuses presented a shear thinning behavior and the viscosity values were in the physiological range: 7.61 ± 0.53 cP for the asymptomatic and 2522 ± 243.3 cP for the symptomatic (**Figure 3**).

**Figure 3.**
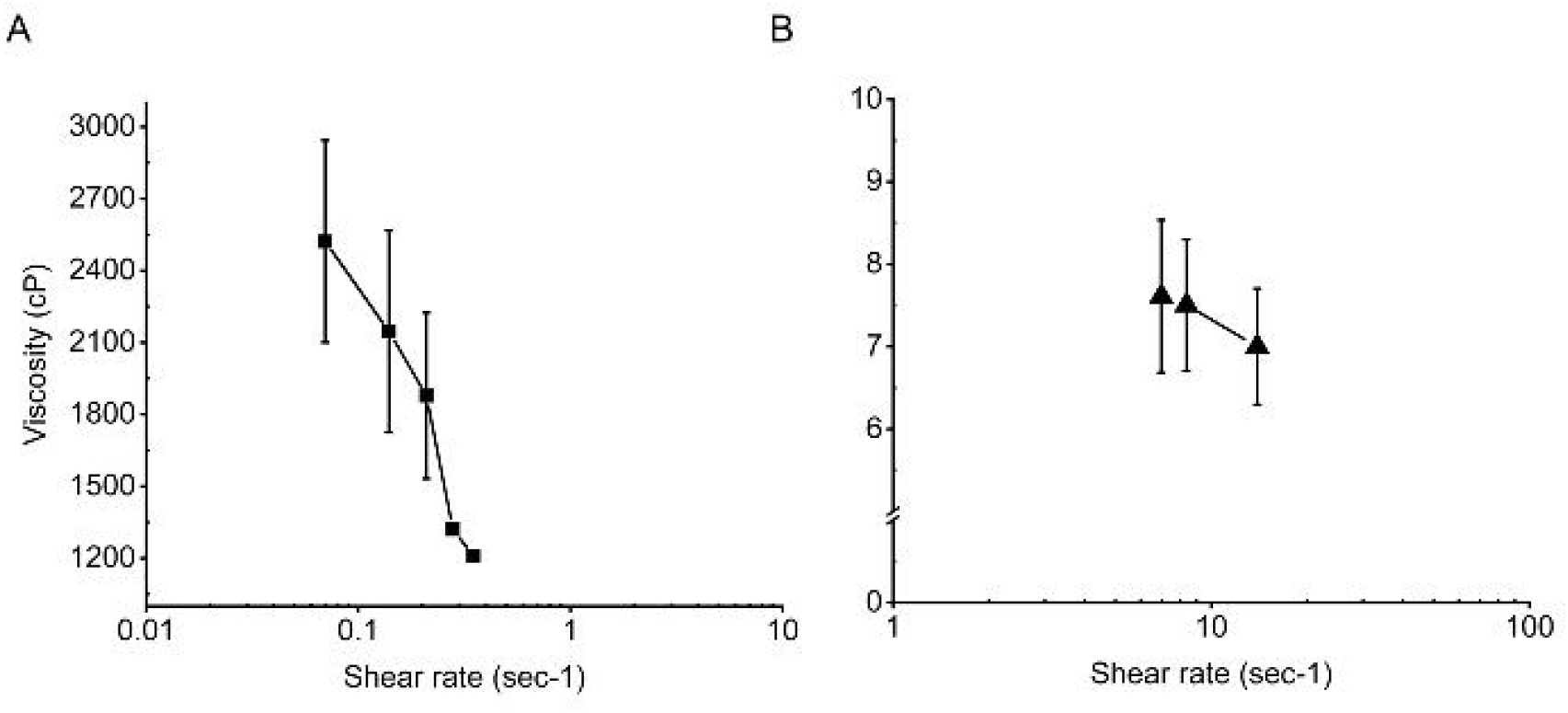
Physical characterization of nasal fluids. Representative viscosity curves showing the effect of shear increment on PEO viscosity at (**A**) 3% w/v (symptomatic, square) and (**B**) 0.5% w/v (asymptomatic, triangle).

### Quantification of swab pick-up and release

The gravimetric analysis was conducted to understand mucus pick-up, expressed as difference in mass, between IM-NP and IM-MT and SF swabs. The tissue-model analysis showed that the SF-NP picked up 1.3 times more 3% w/v PEO than the IM-NP swabs and picked up 4.1 times more 0.5% w/v PEO, while the SF-MT swabs picked up 1.5 times more 3% w/v PEO than the IM-MT and 4.5 times more 0.5% w/v PEO. As a comparison method, we performed the same analysis by dipping the swabs into the same solution in a tube (TUBE method). The TUBE method showed a 2.9 time increase in pick-up of the 3% w/v PEO from IM-NP swabs in comparison to the MODEL collection method, while the SF-NP swab picked up 2.4 times more. The IM-NP swab picked up 5.2 times more 0.5% w/v PEO while the SF-NP swab picked up 1.9 times more 0.5% w/v PEO. The gravimetric analysis concluded that all swabs picked up significantly more mucus in the TUBE method than the MODEL collection method (**Figure 4**).

**Figure 4.**
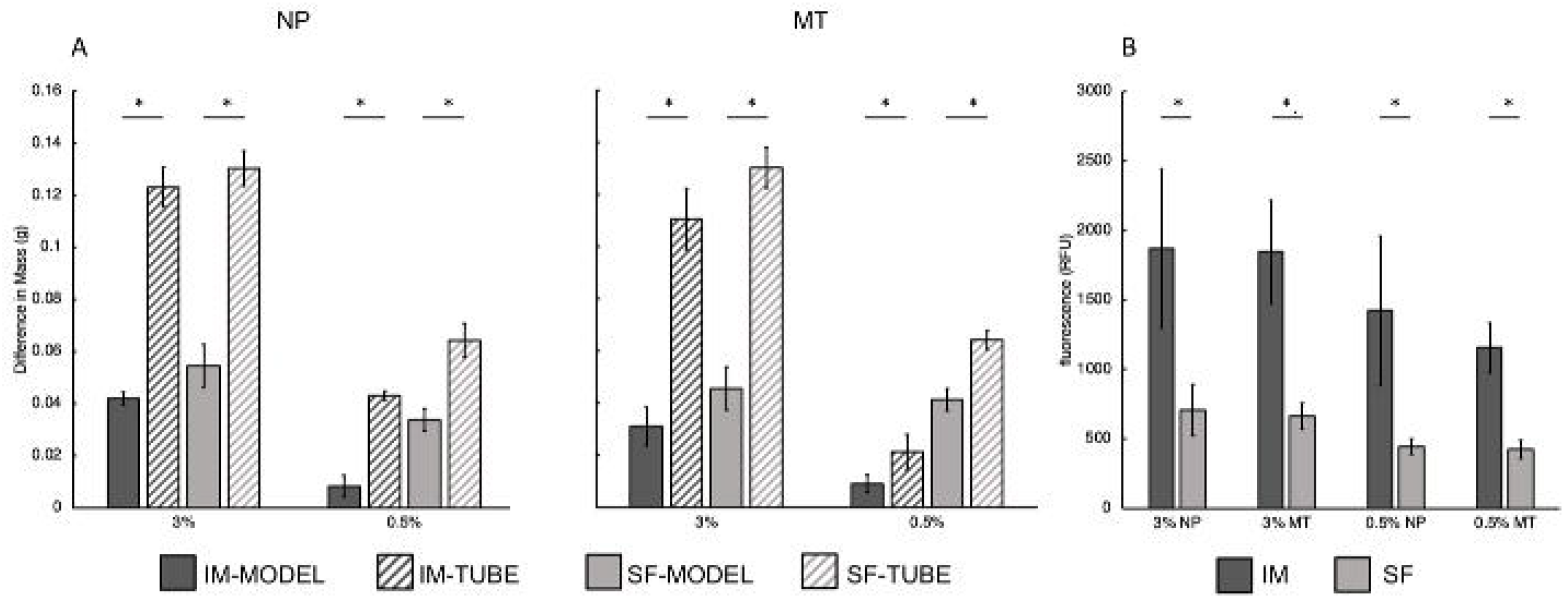
Quantification of swab pick-up and release. (**A**) Gravimetric analysis of IM and SF NP and MT swabs in 3% and 0.5% w/v PEO. The results show the mass pick up of injection molded swabs (IM), and standard flocked swabs (SF) in the tissue model (MODEL) in comparison to the swab dipping standard procedure (TUBE). (**B**) Release quantification of IM and SF NP and MT swabs in 3% and 0.5% w/v PEO loaded with 80% v/v FITC-labeled microparticles. Statistical analysis conducted with t-test, **\***p < 0.05.

The release quantification of FITC-labeled microparticles was performed to efficiently mimic cellular uptake, and subsequently correlate with cellular material release via RT-qPCR analysis. The IM-NP swabs released 2.6 times more microparticles than the SF-swabs in 3% w/v PEO, while in 0.5% w/v PEO the IM-NP swabs released 3.2 times more microparticles. Overall, the IM-NP and IM-MT swabs released statistically significant more microparticles in both asymptomatic and symptomatic conditions in comparison to SF swabs. (**Figure 4**).

### Quantification of swab performance

Bench top validation of swab performance was performed on both IM and SF swabs with the nasal tissue model saturated with synthetic mucus, in symptomatic and asymptomatic conditions, spiked with different loads of SARS-CoV-2 heat inactivated virus, in an effort to encompass clinical variability. The Ct values for all the swabs were compared across the three virus concentrations for both mucus viscosities. Our analysis showed, as expected, that as concentration of virus increased there was also a decrease in Ct values. In fact, there was a 4.48 Ct decrease in the IM-NP swabs in 3% w/v PEO from 10^7^ to 10^5^ copies/mL, and a 4.04 Ct shift in the SF-NP swabs in the same conditions. IM-NP swabs with 10^7^ copies/mL in 0.5% w/v PEO, instead, showed a 5.33 higher Ct value than the 3% w/v PEO Ct. Furthermore, all IM and SF swabs loaded with 10^5^ copies/mL of virus in 0.5% w/v PEO showed a statistical difference between the paired swabs groups in favor of IM swabs. For 10^6^ copies/mL the NP and MT swabs in 0.5% w/v PEO were statistically different, but in favor of the SF swabs. With 10^7^ copies/mL only the IM, and SF-MT swabs were statistically different in 0.5% w/v PEO. For the symptomatic mucus (3% w/v PEO) the MT swabs were statistically different only when they were loaded with 10^6^ or 10^5^ copies/mL, indicating that the IM-MT swabs perform better at lower virus concentration compared to the SF swabs. (Figure 5)

**Figure 5.**
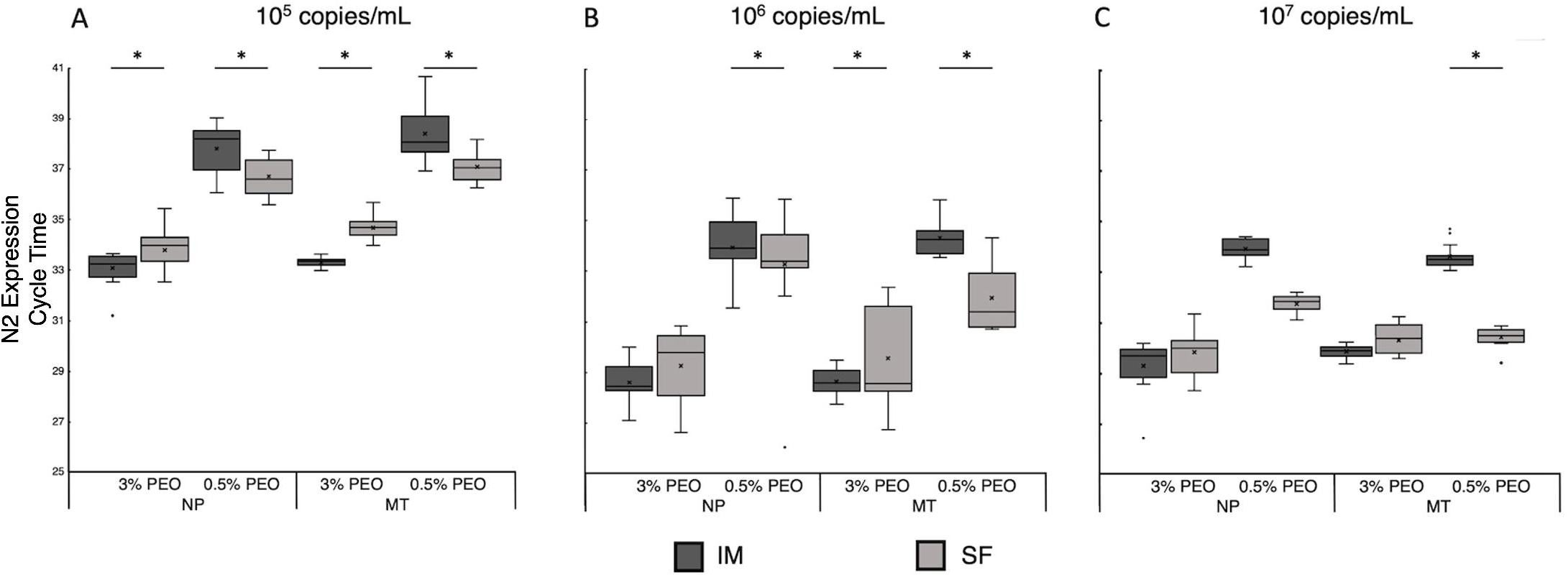
Quantification of swab performance. RT-qPCR quantification of N2 SARS-Cov-2 gene pick up and release for injection molded (IM) and standard flock (SF), nasopharyngeal (NP) and mid-turbinate (MT) swabs validated in a nasal tissue model loaded with symptomatic (3% w/v) and asymptomatic (0.5% w/v) mucus mimicking nasal solutions, spiked with 10^5^(A), 10^6^ (B), and 10^7^ (C) copies/mL of heat-inactivated SARS-Cov-2 virus. * p<0.05

## Discussion

The increasing number of COVID-19 positive cases in the United States led companies to develop new swabs to overcome pandemic associated testing bottlenecks. However, new swabs need to undergo extensive validations prior to reaching the market. To support initial pre-clinical validation, we developed an *in vitro* 3D-printed nasal tissue model, that recapitulates key features of the nasal cavity (i.e., architecture, soft tissue, and mucus viscosity). Alternative strategies aimed to create tissue models that mimicked the paranasal sinuses and skull and were flexible enough to guide surgeons in their preoperative practice simulations ^20^. Although useful, those models were meant to be a valuable tool for surgeons and not for testing and research purposes. For this reason, we have developed a model that could efficiently and safely support research and development stage of medical devices to streamline the validation of new swab prototypes and increase its clinical relevancy before clinical studies.

Our initial anterior nasal passage model consisted of a silicone tubing lined with a cellulose sponge ^5^, with sizes compatible with the human nostril ^19, 21^. This model was subsequently modified with the proposed 3D-printed tissue model, to replicate the entire structure of the mid-turbinate and nasopharyngeal walls of the nasal cavity. The degree of precision achieved in the reproduction of the nasal cavity in our 3D-model was accomplished by averaging the computed tomography (CT) scans of 30 healthy patients’ nasal cavities provided by the Aerosol Research Lab at Carleton University in Ottawa-Canada ^14^. The model was then lined with a silk sponge, as a replacement for the cellulose sponge to better mimic soft tissue mechanical properties ^22^, and with synthetic mucus fluids to resemble both symptomatic and asymptomatic fluid viscosities. Silk protein was chosen because of its structural and mechanical properties and inertness. Silk is a versatile, biocompatible, and biodegradable material with tunable mechanical properties and is extensively used in tissue engineering for mimicking soft and high-strength human tissues ^23-26^. In our model, silk was used in a sponge format to replicate the soft architecture of the nasal tissue with a controlled pore size. Lastly, to mimic the nasal mucus, we utilized PEO, a hydrophilic polymer with physical and mechanical properties that can be tuned based on its molecular weight ^27^. Due to its viscoelastic properties, PEO at different concentrations creates a range of viscous solutions that can be used to mimic physiological mucus ^28^. Another advantage to using PEO is its compatibility with biomolecular assays; in fact, PEO does not interfere with RT-qPCR amplification at low viscosities compared to other viscous body fluids ^29^. To match the viscosity of the nasal fluid in asymptomatic and symptomatic conditions, we tested several PEO concentrations finding that 0.5% and 3% w/v were compatible with the physiological range of nasal fluid viscosities ^17^. In general, human mucus viscoelasticity is characterized by a shear thinning behavior with a viscosity range between 10 and 10^6^ cP ^17^. Furthermore, low, and high mucus viscosities have been associated with asymptomatic and symptomatic nasal mucus viscosities (∼13 and 1400 cP) in artificial mucus compositions ^30^. Our synthetic mucus formulation confirmed the shear thinning behavior ^18^ and matched the viscosity range for both nasal fluid conditions (**Figure 3**).

To support our model as a suitable tool for swab validation, we developed and performed several assays to assess swab pick up and release efficiencies and then evaluate data agreement against available literature. In addition, an *in vitro* tissue model would provide more controlled experimental conditions, in comparison to clinical studies that have shown greater variability, arising from differences in sampling methods, nasal cavity structure, nasal fluid viscosity, and other conditions that vary from patient to patient and season to season ^3-4^. Moreover, the disadvantages of relying on clinical trials for initial swab validation are also the bureaucratic aspects (i.e., recruitment, regulatory requirements, and cost). Thus, the tissue model would be a great tool to support initial research and development explorations with clinical relevancy for swab design and optimization. Data from clinical trials showed that IM swabs perform similarly to SF swabs and those results are comparable with our findings ^3-5^. In fact, the IM2 IM swabs had an agreement of 96% with the FLOQ standard flocked swab during their clinical trials ^3^, as also supported by our findings with the *in vitro* tissue model.

We initially quantified swab pick up in our model via gravimetric analysis and subsequently quantified viral release via RT-qPCR. Current methodologies simulate the specimen collection by dipping the swab into a spiked COVID-19 negative nasal fluid or water and estimating the swab pick up by measuring, pre and post, its weight or volume (TUBE method). However, such analyses can lead to misleading results when analyzing the data ^1, 4^, because they replicate neither the nasal architecture and physiological fluids nor the actual collection procedure; furthermore, when comparing swab typologies, data in literature reported that SF swabs dipped in water pick up 10.7 times more water than the IM swab due to their difference in geometry, material and device fabrication ^4^. Another discrepancy in the TUBE method as a workflow to correctly assess pick up and release when using contrived samples is the viscosity of the solution used. For example, a PurFlock Ultra flocked swab picked up 6.3·10^4^ copies of viral material from a tube method ^1^, while a flocked swab picked up 1.6·10^4^ copies from our model with 0.5% w/v PEO. Our analysis in fact showed that the standard NP picked up 1.9 times more mucus (0.5% w/v PEO) from the TUBE method than the MODEL method. This suggests that the dipping TUBE model allows for a much greater absorption of liquid, and therefore more viral material, in comparison to a standard swabbing procedure, introducing artifacts in the data collection (**Figure 4**). This supports the importance in replicating the native architecture in the validation process.

On the other hand, a release quantification analysis needs to be performed in order to obtain reliable and consistent molecular data for the virus detection. This type of analysis has been done in the past with different microorganisms, where contrived biological samples were dispensed by pipetting onto swabs, eluted in a buffer, and then processed for RT-qPCR ^31^. This method, however, does not actually replicate the specimen collection and release process that, again, are dependent on the swab pick-up features as well as the anatomical structure and geometry of the patient cavities. Our data, in fact showed that the IM-NP swabs released more microparticles compared to the standard swabs which can be attributed to their geometry and structural properties. In fact, SF swabs do not release as many microparticles as the IM due to their nylon fibers which are meant for maximum absorbance and high retention. On the contrary, the hydrophobic plastic nature of the IM swabs allows them to release most of the sample they collect ^4^ (**Figure 4**).

As a final test, we concluded the validation of our model by performing RT-qPCR. We initially tested the limit of detection of the model by spinking the synthetic nasal mucus with 10^5^, 10^6^ and 10^7^ copies/mL of heat inactivated SARS-CoV-2 virus. In general, clinical studies typically compare Ct values to a control since the viral load is unknown during the specimen collection and diagnostic process. Our analysis demonstrated that when the model is spiked with decreasing concentration of COVID-19 virus, the Ct values were acceptable among all the conditions except for 10^5^ in 0.5% w/v (38 Ct); however, according to the World Health Organization (WHO) a Ct value between 37 and 40 is at the acceptance limit ^32^. The same behavior was evident also when the virus was spiked at 10^7^ copies/mL in the 0.5% w/v PEO in which there was a difference of 5 cycles for IM-NP and 4 cycles for SF-NP, compared to 3% w/v PEO, even if the values are in the acceptance range (Ct≤37). Those differences must be caused by the lower viscosity of the mucus which causes a major dispersion of the virus in the sponge and, consequently, a lower pickup or release by the swab (**Figures 5** and **S1**). In addition, IM swabs confirmed comparable performance to SF swabs for higher viral loads, while they outperform SF swabs in all symptomatic conditions. This supports the importance of replicating the physical properties of the native tissue in the validation process.

## Conclusion

Global shortage of collection specimen swabs has been among the several bottlenecks in COVID-19 testing, during the 2020 pandemic. Several companies have created new injection molded swabs that can be mass produced quickly and cost efficiently. To validate these swabs, we have developed an *in vitro* tissue model of the human nasal cavity. This model accurately mimics the architecture and structure of the cavity and is lined with silk sponges to resemble the nasal soft tissue. An artificial mucus was also developed from PEO to replicate two different physiological conditions, asymptomatic and symptomatic nasal fluid viscosities. This model was used to validate a new injection molded swab and to provide comparable RT-qPCR results to a standard flocked swab, showing the importance of replicating physical, and structural features of the native tissue as part of the validation process. We are confident that this model will be critical to support pre-clinical validation stages, by efficiently saving time and money during medical device validation.

## Data Availability

All data produced in the present study are available upon reasonable request to the authors

## ASSOCIATED CONTENT

**Supp. Figure S1.**
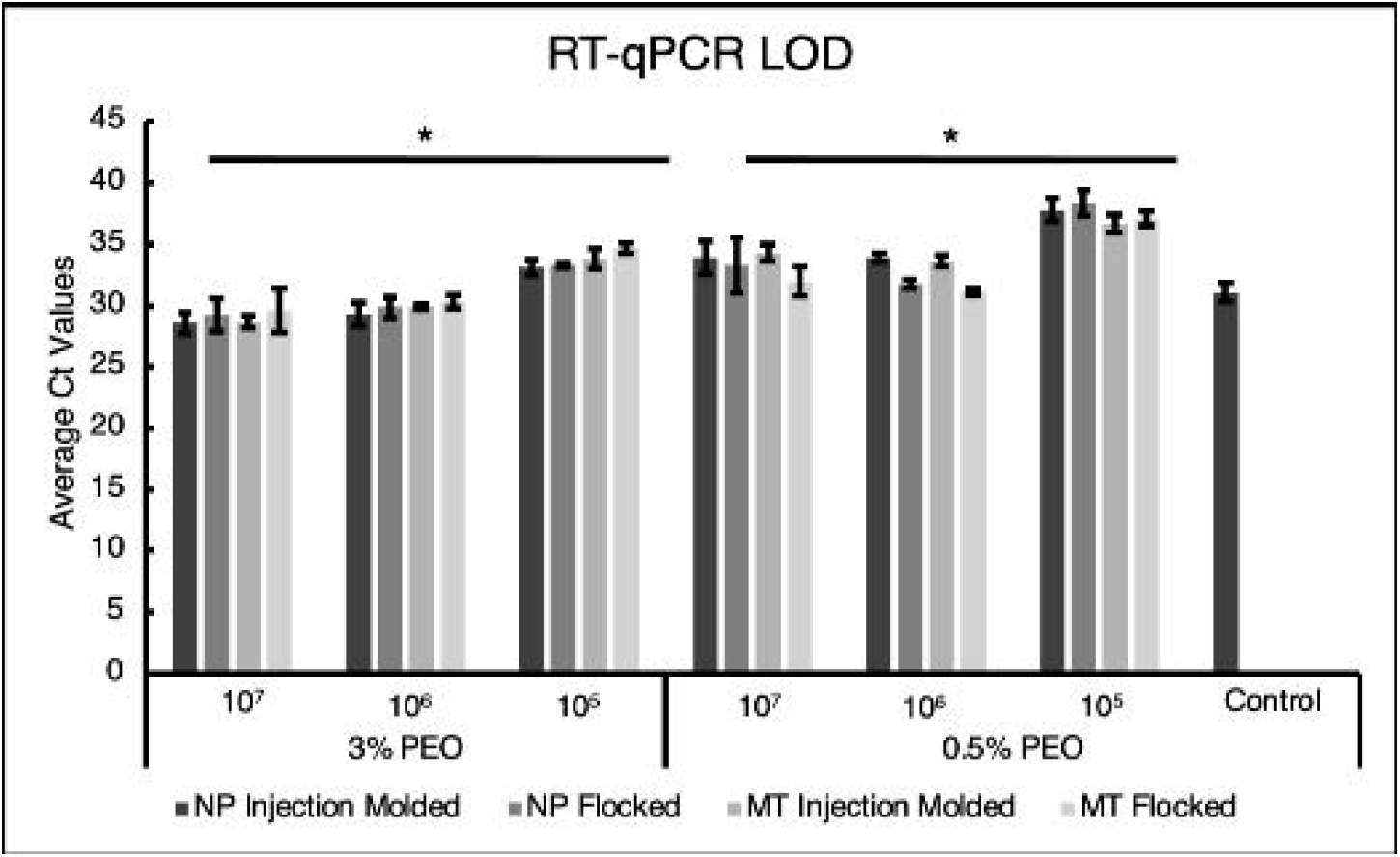
Average Ct values for all concentrations of SARS-CoV-2 virus, PEO, and types of swabs including a RT-qPCR positive control. The control consists of 10^6^ copies/mL of virus in nuclease free water (VWR Life Sciences, Pennsylvania). Statistical analysis was done with ANOVA single factor, * p < 0.05

## Author Contributions

The manuscript was written through contributions of all authors. All authors have given approval to the final version of the manuscript.

## ACKNOWLEDGMENT

We would like to thank M2D2 for 3D printing access, Professor F. Omenetto and graduate student N. A. Ostrovsky-Snider (Tufts University) for helping us with the viscosity measurements. We would also like to thank the Aerosol Research Lab at Carleton University in Ottawa, Canada for the .stl file of the nasal cavity model.

## ABBREVIATIONS

IM: injection molded swab
SF: standard flocked swab
MT: mid-turbinate swab
NP: nasopharyngeal swab
PEO: Poly (ethylene oxide)

